# Assessing Neuropsychiatric Symptoms in Long COVID: A Retrospective Cohort Study from a South Texas Long COVID Clinic

**DOI:** 10.1101/2024.11.03.24316669

**Authors:** Anne Marie Wells, Summer Rolin, Barbara Robles-Ramamurthy, Gabriela Gibson-Lopez, Martin Goros, Jonathan A Gelfond, Stephen Gelfond, Philip Balfanz, Melissa Deuter, Donald McGeary, Monica Verduzco-Gutierrez

**Affiliations:** South Texas Medical Scientist Training Program, UT Health San Antonio, San Antonio, TX, USA; Department of Pharmacology, UT Health San Antonio, San Antonio, TX, USA; Department of Rehabilitation Medicine, UT Health San Antonio, TX, USA; Department of Family & Community Medicine, UT Health San Antonio, San Antonio, TX, USA; Department of Psychiatry & Behavioral Health Sciences, UT Health San Antonio, San Antonio, TX, USA; South Texas Psychiatry Practice-Based Research Network (PBRN), San Antonio, TX, USA; Department of Population Health Sciences, UT Health San Antonio, San Antonio, TX, USA

**Keywords:** PASC, Long COVID, PTSD, stress, depression, anxiety, PCL-5, GAD-7, PHQ-9

## Abstract

Long COVID, previously known as Post-Acute Sequelae of SARS-CoV-2 (PASC), refers to prolonged symptoms or diagnosable conditions following COVID-19 infection. The neuropsychiatric profile of Long COVID patients remains ambiguous. This study aimed to assess neuropsychiatric symptoms in a retrospective cohort of Long COVID patients (N = 162) at a Rehabilitation Medicine clinic in South Texas. Clinical data from patient records were used to calculate a Symptom Score, and screening tools for stress/PTSD (PCL-5), depression (PHQ-9), anxiety (GAD-7), and quality of life (SWL) were employed to evaluate if Long COVID duration and severity could predict neuropsychiatric outcomes. The majority were female (71%) and Hispanics (53%) who presented for treatment of Long COVID symptoms during the study period, including fatigue (93%), coughing/shortness of breath (81%), fever (67%), anosmia (58%), ageusia (54%), and weight loss (56%). A minority of participants were hospitalized (N = 49) or required ventilator support (N = 5) during acute infection. There was a high burden of neuropsychiatric symptoms, including subjective cognitive impairment (79%), headache (74%), and insomnia (58%). Symptom Score (median = 9, IQR [8,11]) was significantly correlated with increased depression (PHQ-9; p < 0.05), anxiety (GAD-7; p < 0.05) and elevated stress/PTSD (PCL-5; p < 0.05) symptoms. Long COVID patients taking stimulants or mood stabilizers had higher GAD-7 (p < 0.031, p < 0.035) and PHQ-9 (p < 0.034, p < 0.009) scores but not PCL-5 scores. Importantly, duration of Long COVID symptomatology also did not predict PCL-5 scores. No patient factors (e.g., sex, age, BMI, ethnicity) mediated Symptom Score. Nonetheless, historically marginalized groups, such as women and Hispanics, have been disproportionately affected by COVID-19. This study is the first to utilize validated screening tools to determine the presence and severity of neuropsychiatric symptoms in Long COVID patients. These findings may guide clinical management and future research on Long COVID, especially in historically excluded populations.

**Scope Statement:** We enthusiastically submit our Original Research article, entitled “**Assessing Neuropsychiatric Symptoms in Long COVID: A Retrospective Cohort Study from a South Texas Long COVID Clinic**” for consideration for publication in the journal Frontiers in Neurology. We believe the scope of our article aligns well with the scope and aim of the journal’s Neurorehabilitation Section.

Long COVID is a debilitating neurological disorder with prominent and enduring cognitive and psychological impact. This study sought to characterize Long COVID symptoms from a cohort of patients at a Rehabilitation Medicine/Long COVID clinic in Southwest Texas. We stratified symptoms using validated psychiatric evaluation tools (e.g., PCL-5, GAD-7, PHQ-9, SWL) to determine if and to what extent psychiatric comorbidity exacerbated Long COVID symptoms. Our findings suggest that a Long COVID patient’s depression, anxiety, and stress/post traumatic stress scores are highly correlated with other neurological symptoms. We advance the implementation of a Long COVID “Symptom Score”, as well as the use of validated screening instruments to identify psychiatric features of Long COVID with the goal of maximizing life satisfaction and function over the course of treatment.

## Introduction

The impact of the SARS-CoV-2 virus, which causes acute COVID-19 infection, is an ongoing public health concern in the United States (US) that is here to stay (1). Continued study of the multifaceted impact on health and longevity is essential to advance the development of effective therapeutic and prevention strategies. Most individuals recover from COVID-19 infection within 5-20 days, depending on severity of symptoms (2,3); yet, a recent meta-analysis (4) estimated 31-69% of COVID-19 patients endure ongoing, relapsing and remitting, or progressive symptoms beyond 30 days of primary infection. The resulting syndrome is known as Post-Acute Sequelae of SARS-CoV-2 (PASC) (5) or Long COVID. Thus, Long COVID is now defined by the National Academy of Sciences, Engineering, and Medicine as an infection-associated chronic condition that occurs after SARS-CoV-2 infection and is present for at least 3 months as a continuous, relapsing and remitting, or progressive disease state (6). Aggregated Long COVID symptomatology (5,7–9) has been tracked in both small (10) and large (9,11,12) longitudinal cohort studies. In addition to continued symptoms of the acute infection (e.g., coughing, shortness of breath, loss of taste and smell), all studies to date have identified a common clustering of symptoms primarily affecting neurologic or neuropsychiatric systems (4,5,9–12). These symptoms include chronic fatigue, cognitive impairment (“brain fog”), headache, pain syndromes, anxiety and depression. Given the neurovirulent profile of SARS-CoV-2 (13), one might predict that the neuropsychiatric burden of Long COVID contributes significantly to complex treatment need and delayed recovery of affected patients. In fact, most patients experience reduced health-related quality of life (14) and Long COVID neuropsychiatric cluster symptoms for at least 6 months and upwards of 3 years out from primary infection (9). Thus, Long COVID poses a significant and chronic disruption to patient lives, as demonstrated by increased disability and economic burden reported in this population (15).

Known risk factors (9,15–17) for Long COVID include those that predicate more severe COVID-19 infection, such as obesity, age, premorbid metabolic or cardiovascular conditions -- and, critically, healthcare equity and access. Historically excluded groups in the US, like women and Hispanics, have been disproportionately impacted by the COVID-19 pandemic (15–20). A recent multi-site study following over 12,000 patients (73% female) found that female sex was significantly associated with higher risk of Long COVID (21). While there are currently no large cohort studies following Hispanic-Americans, one study (22) sampling adults in Mexico found at least 5 persistent Long COVID symptoms in over half of participants (N = 192), with 360-day persistence probability of 0.78. Strikingly, the largest cohorts following the natural history of Long COVID in the US have disproportionately sampled Caucasian men (9,11). Thus, there may be significant gaps in our understanding of how various patient factors may impact the complexity and duration of Long COVID symptom clusters.

There are no cures nor adequate treatments for Long COVID apart from symptom management, perhaps due the complex, syndromic nature of the disorder. Importantly, studies published to date have relied entirely on self-reported neuropsychiatric symptoms without the support of validated screening tools that could facilitate uncovering the etiology of neuropsychiatric symptoms. There is a high degree of clinical utility in defining quantifiable metrics to expediate diagnosis and treatment of the neuropsychiatric burden of Long COVID.

We hypothesized that while chronic stress and anxiety do not directly facilitate the development of Long COVID, the presence and severity of neuropsychiatric symptoms should track with the core symptomatology and drive symptom complexity. Thus, we sought to characterize Long COVID symptoms in a cohort of patients (N = 162; 71% female, 53% Hispanic, median BMI = 30 [obese]) who presented in a Rehabilitation Medicine clinic in South Texas. We applied a proof-of-concept methodology to rapidly screen for major clusters of Long COVID symptomatology (i.e., “Symptom Score”) that can be employed with a battery of widely used screening tools for comorbid stress, anxiety, and mood disorder neuropsychiatric symptoms. Our findings point to the high burden of fatigue, cognitive impairment, and anxiety measures in Long COVID patients that exacerbate the core Long COVID symptom profile. The associated impact on quality of life and risk of worsening psychiatric conditions demands greater attention to neuropsychiatric symptoms in the management of Long COVID patients, particularly in marginalized groups.

## Results

### Long COVID patients report high rates of fatigue with neuropsychiatric symptomatology

We characterized the self-reported symptom profile in Long COVID patients (N = 162), defined as symptoms or conditions present for at least 30 days after acute COVID-19 infection. Symptoms and conditions considered in this study were based on major categories determined by previous studies(5,9,11). The top-ranked symptoms within this cohort are summarized in Figure 1. Of note, 89% of the sample received an initial COVID-19 infection diagnosis by positive test (Fig 1). The distribution of approximate time elapsed since COVID-19 infection was approximately normal (Fig S1A). Of note, all patients received initial COVID-19 diagnosis within one of the major peaks of the COVID-19 pandemic (Fig S2).

**Figure 1.**
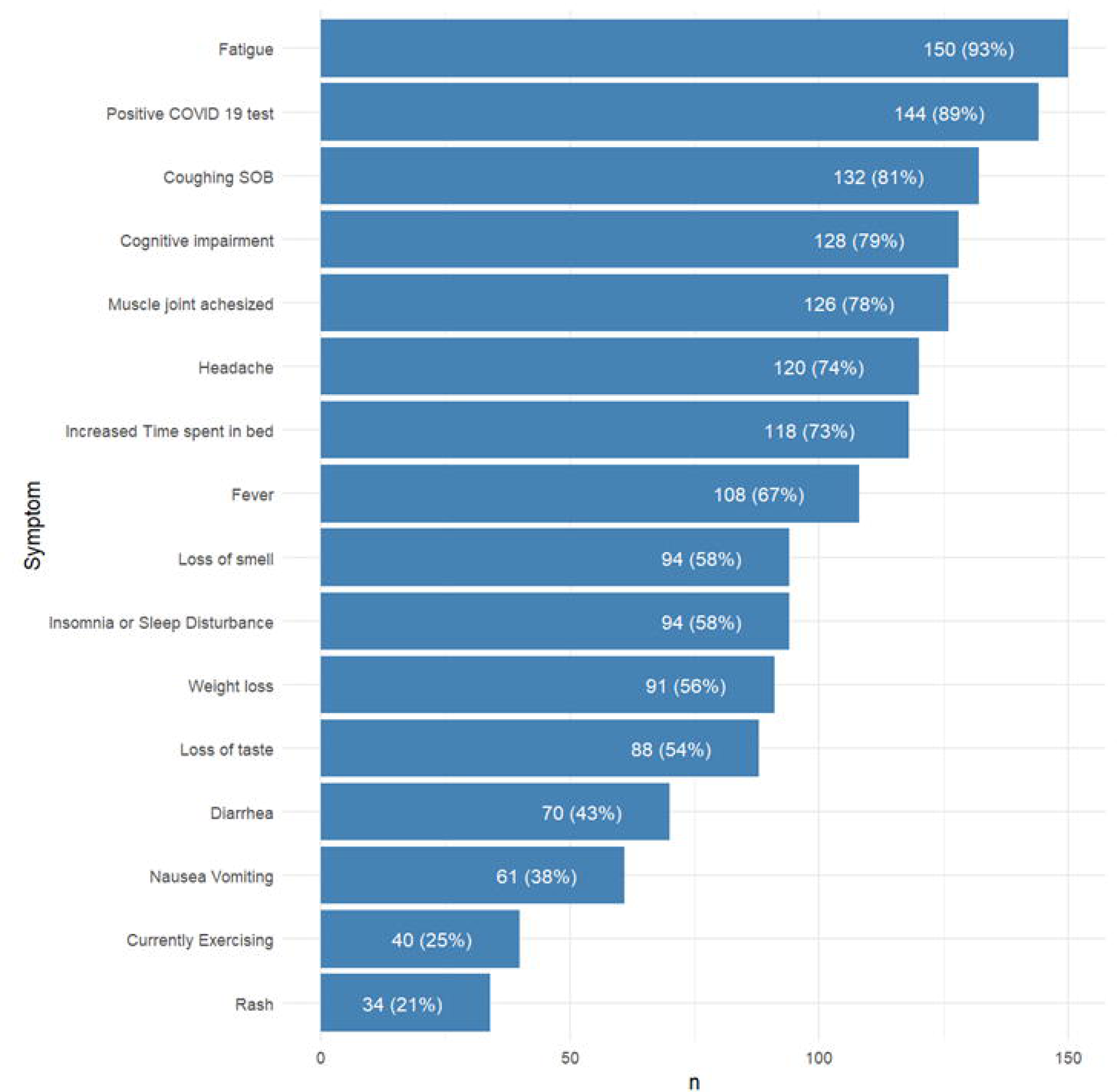
Summary of Symptom Score Components and incidence (%) in analytical sample. We collected self-reported data on PASC symptomatology across all major systems as previously described (Bowe et al 2023; Cai et al 2024). Symptoms were ranked by incidence (% of patients who reported symptom) in our sample. We then calculated a PASC Symptom Score based on the total number of comorbid symptoms present in a patient based on the top 16 most common symptoms, summarized here. Of note, the most common PASC symptoms noted by our sample was fatigue (93%). There was a high burden of symptoms classically related to acute COVID-19 infection (coughing, shortness of breath [SOB], loss of smell and taste) as well as subjective neuropsychiatric symptoms (cognitive impairment = 79%, muscle/joint pain = 78%, insomnia/sleep disturbance = 58%).

The most common symptom reported in our cohort was fatigue (93%). At time of assessment, only 25% were currently exercising. Patients commonly reported myriad symptoms commonly associated with Long COVID fatigue(23), such as gastrointestinal (diarrhea [43%]), nausea/vomiting [38%], weight loss [58%]) and autoimmune (rash [21%]) symptoms. Most patients also reported a prolonged cluster of symptoms of acute COVID-19 infection, including coughing or shortness of breath (81%), fever (67%), loss of smell (58%), and loss of taste (54%).

The burden of neuropsychiatric symptoms in this cohort was striking. A substantial number of patients reported subjective cognitive impairment (79%), headache (74%), increased time spent in bed (73%), insomnia/sleep disturbance (58%). Many patients also reported new onset of muscle and/or joint pain (78%).

Importantly, we noted that very few patients with Long COVID symptoms in this cohort reported being hospitalized (N = 49) or requiring a ventilator (N = 5) during initial COVID-19 infection. Moreover, few required oxygen therapy or sustained cardiac damage because of COVID-19 infection. Thus, our cohort largely represents a patient group with Long COVID symptomatology following non-severe primary COVID-19 infection.

### Patient factors do not predict Long COVID Symptom Score

We next sought to determine if patient factors predicted Long COVID symptom burden (Table 1). To this end, we ranked the incidences of self-reported symptoms or conditions present in our cohort. We then summed the total number of symptoms reported from the top 16-ranked symptoms for each patient as a Symptom Score. The median Symptom Score was 9 (IQR [8, 11]). We analyzed the relatedness of Symptom Scores across the cohort to patient factors. We also sought to determine if there were significant differences in patient factors between two subgroups divided by the median Symptom Score. We considered various patient factors, including biological sex, age, body mass index (BMI), race and ethnicity.

**Table 1.** Summary of patient factors and prediction of Long COVID Symptom Score.

| Patient Factor | PASC Symptom Score |  |  | p-value<sup>2</sup> |
| --- | --- | --- | --- | --- |
|  | Overall, N = 162<sup>1</sup> | > Median, N = 78<sup>1</sup> | ≤ Median, N = 84<sup>1</sup> |  |
| Symptom Score | 9 (8, 11) | 11 (10, 12) | 8 (6, 8) | <0.001 |
| Sex |  |  |  | 0.2 |
| Female | 117 (73%) | 60 (78%) | 57 (68%) |  |
| Male | 44 (27%) | 17 (22%) | 27 (32%) |  |
| (Missing) | 1 | 1 | 0 |  |
| Age at Evaluation | 44 (35, 54) | 44 (36, 52) | 46 (34, 56) | 0.4 |
| BMI | 30 (26, 36) | 30 (26, 35) | 29 (26, 36) | 0.7 |
| (Missing) | 22 | 12 | 10 |  |
| Race/Ethnicity<sup>3</sup> |  |  |  | 0.8 |
| Black or African American | 6 (3.9%) | 4 (5.3%) | 2 (2.6%) |  |
| White | 61 (40%) | 29 (39%) | 32 (42%) |  |
| Hispanic | 78 (51%) | 38 (51%) | 40 (52%) |  |
| Other | 7 (4.6%) | 4 (5.3%) | 3 (3.9%) |  |
| (Missing) | 10 | 3 | 7 |  |
| Time since infection (Days) | 190 (83, 278) | 218 (83, 295) | 169 (95, 264) | 0.4 |
| (Missing) | 5 | 0 | 5 |  |
| Time since infection < 30 Days | 8 (5.1%) | 1 (1.3%) | 7 (8.9%) | 0.063 |
| (Missing) | 5 | 0 | 5 |  |

<sup>1</sup> Median (IQR); N (%)
<sup>2</sup> Wilcoxon rank sum test; Pearson's Chi-squared test; Fisher's exact test (categorical variables)
<sup>3</sup> Other includes: American Indian Alaska Native, Asian. Unknown set to Missing

More female (N = 117, 73%) than male (N = 44, 27%) patients presented for evaluation for Long COVID symptoms during the study period. There was a higher proportion of females with above median symptom score compared to males, but this difference did not reach statistical significance (78% vs 68%, *p* = 0.2). Thus, despite significantly more females in our sample, there was no contribution of biological sex to Symptom Score. Moreover, age (median= 44, IQR [35, 54]) and BMI (median = 30, IQR [26,36]) also failed to predict Symptom Score (*p* = 0.4, 0.7, respectively). Likely due to the geographic location of this study, most patients identified as Hispanic (53%). Yet, no race or ethnicity identified in this cohort were associated with Symptom Score (*p* = 0.8).

### Stress, anxiety, and satisfaction with life vary amongst Long COVID patients

Given the high contribution of neuropsychiatric symptoms to Symptom Score and disease burden, we sought to determine if the additional psychiatric symptoms could predict Symptom Score. In a first, we administered validated screening tools to assess the presence and severity of stress rising to the level of post-traumatic stress disorder (PTSD; PCL-5), anxiety (GAD-7), depression (PHQ-9), and general satisfaction with life (SWL).

The categorical score results across these screening tools are summarized in Table 2. PCL-5 score revealed 44% of the sample lived with stress-related symptoms that rise to the level of PTSD. We were unable to analyze this finding considering the index (inciting) traumatic event, and thus cannot attribute symptoms to Long COVID symptoms or COVID-19 infection specifically. GAD-7 scores demonstrated 69% of Long COVID patients demonstrated at least mild anxiety symptoms with 18% affected by severe anxiety. Most (74%) patients screened for depression with PHQ-9 had symptoms characterized by at least mild depression with 28% experiencing moderately severe to severe symptoms. Of note, approximately 41% of the cohort was slightly to extremely dissatisfied with life.

**Table 2.** Categorical score results of screening tools for stress (PCL-5), anxiety (GAD-7), and depression (PHQ-9) symptoms, and satisfaction with life (SWL) in Long COVID patients.

| **PCL-5 Score (N = 162)** | **N (%)** |
| --- | --- |
| Not elevated | 90 (56%) |
| Significantly elevated trauma symptoms of PTSD | 72 (44%) |
| **GAD-7 Score (N = 143)** | **N (%)** |
| No Significant Symptoms | 44 (31%) |
| Mild | 38 (27%) |
| Moderate | 35 (24%) |
| Severe | 26 (18%) |
| (Missing) | 19 |
| **PHQ-9 Score (N = 146)** | **N (%)** |
| Minimal Depression | 38 (26%) |
| Mild Depression | 32 (22%) |
| Moderate Depression | 34 (23%) |
| Moderately Severe Depression | 21 (14%) |
| Severe Depression | 21 (14%) |
| (Missing) | 16 |
| **SWL Score (N = 147)** | **N (%)** |
| Dissatisfied | 11 (7.5%) |
| Extremely dissatisfied | 14 (9.5%) |
| Extremely satisfied | 18 (12%) |
| Neutral | 4 (2.7%) |
| Satisfied | 39 (27%) |
| Slightly dissatisfied | 35 (24%) |
| Slightly satisfied | 26 (18%) |
| (Missing) | 15 |

We then sought to determine if Symptom Score informed stress and anxiety levels in this cohort by calculating Spearman rank correlations between each measure (Fig 2). PCL-5, GAD-7, and PHQ-9 scores were all mutually associated. Symptom Score was positively associated with PCL-5 (r = 0.25, *p* < 0.05), GAD-7 (r = 0.29, *p* < 0.05), and PHQ-9 (r = 0.25, *p* < 0.05) scores. Thus, the more complex Long COVID symptomatology, the worse stress, anxiety and depression symptoms a patient reported. We refined this analysis to examine how each of the 20 items of the PCL-5 correlated with individual Long COVID symptoms that comprise the Symptom Score (Fig 3). Nearly all items on the PCL-5 were positively and significantly correlated with insomnia or sleep disturbance (17/20 items, *p* < 0.05).

**Figure 2.**
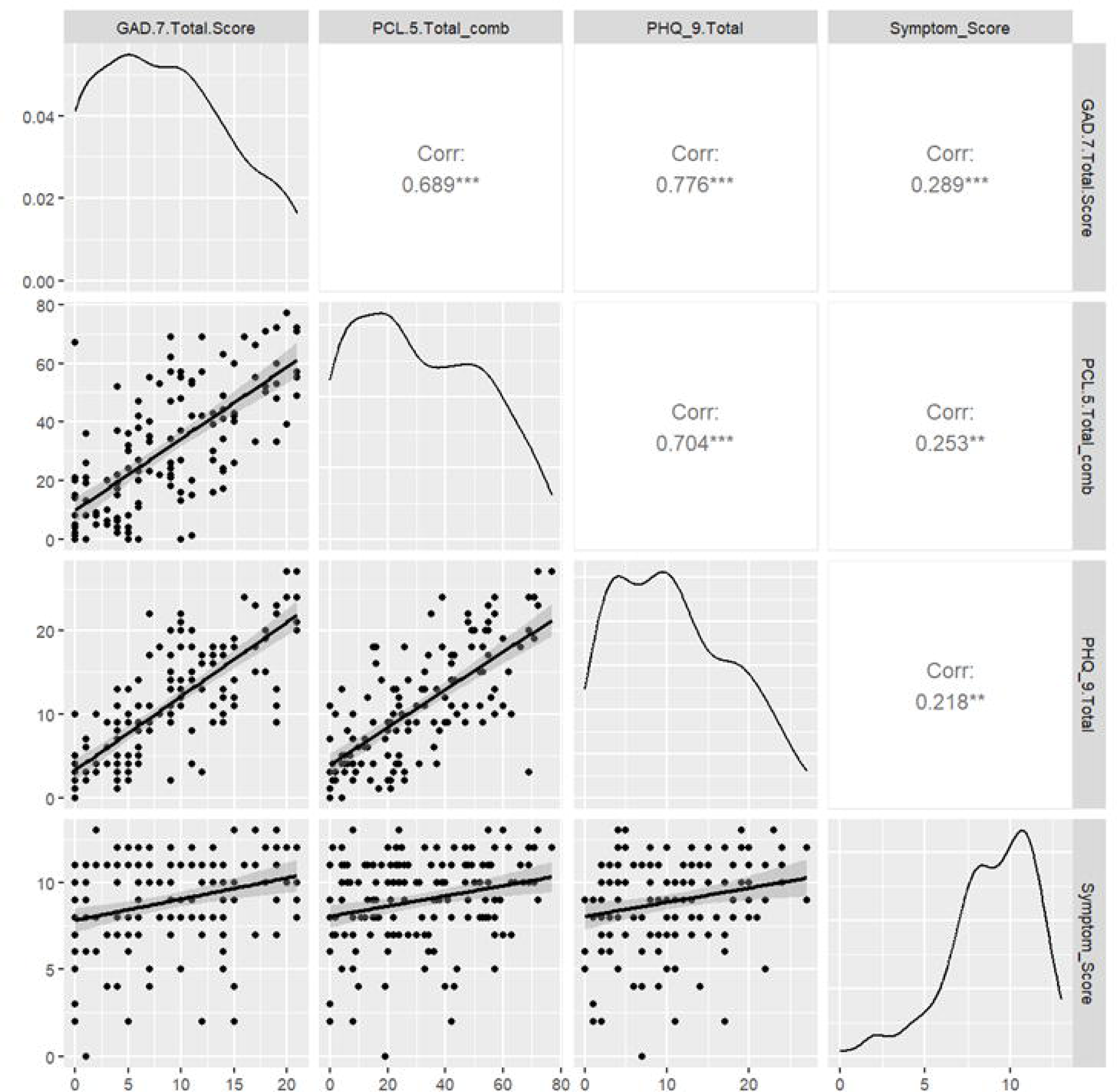
Correlation of Symptom Score with measures of depression (PHQ-9), stress (PCL-5), and anxiety (GAD-7) symptoms. We assessed whether PASC Symptom Scores were correlated with the presence of depression, stress, or anxiety symptomatology as determined by validated screening tools. Spearman rank correlations were computed for each pairwise combination of PASC Symptom Score and PCL-5 or GAD-7 scores. PHQ-9, PCL-5, and GAD-7 scores were mutually associated. PASC Symptom Score was positively associated with PHQ-9 (0.218, *p* < 0.05), GAD-7 (r = 0.29, *p* < 0.05) and PCL-5 (r = 0.25, *p* < 0.05).

**Figure 3.**
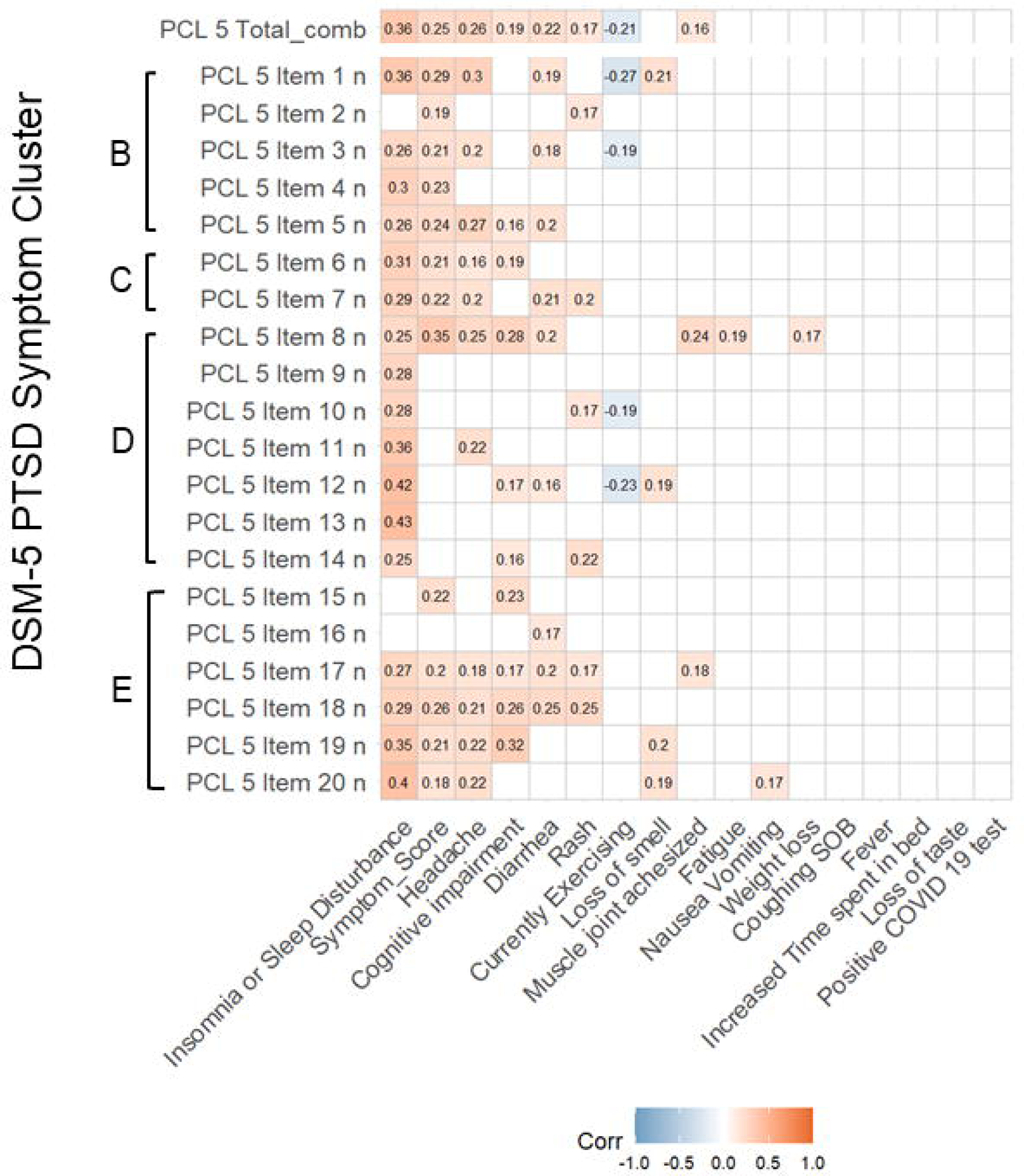
Correlation of individual PCL-5 items with Long COVID Symptoms. Spearman rank correlations were calculated for the intersection of the score of each item, reflecting frequency of stress and PSTD-related symptoms by item of the PCL-5 and the PASC symptoms used to calculate PASC Symptom Score. We contextualize the PCL-5 item scores with their respective symptom clusters for Post-Traumatic Stress Disorder according to the DSM-5, designated to the left of each PCL-5 item: Cluster B (The traumatic event is persistently experienced), Cluster C (Avoidance of trauma-related stimuli after trauma), Cluster D (Negative thoughts or feelings began or worsened after the trauma), Cluster E (Trama-related arousal and reactivity that began or worsened after the trauma. Correlations are given for all findings with *p* < 0.05.

Importantly, individual item correlations do not vary by PTSD Symptom Cluster (B, C, D, E) as identified in the DSM-5. Headache (11/20 items, *p* < 0.05) and subjective cognitive impairment (9/20, *p* < 0.05) were also significantly positively correlated with many items. Of note, whether the patient was currently exercising at time of assessment was negatively correlated with 5 items (*p* < 0.05). The item most positively correlated with the greatest number of symptoms was item 8 *(p* < 0.05): “In the past month, how much were you bothered by: Trouble remembering important parts of the stressful experience?”

### Pharmacotherapy may moderate stress and anxiety-related Long COVID symptoms

We sought to determine if any ongoing pharmacotherapy with Long COVID symptoms could explain the presence of stress and anxiety symptoms in our cohort. To this end, we performed a limited scope analysis by reviewing current prescriptions reported by each patient at the time of evaluation for Long COVID. Importantly, all medications reported were prescribed prior to the intake appointment without knowledge of prescription initiation or purpose of medication. We then correlated PHQ-9, GAD-7 and PCL-5 scores with classes of medications reported taking at the time of Long COVID evaluation. Of note, 41% of patients take medications related to respiratory care well after initial COVID-19 infection (Table S1). Although PHQ-9, PCL-5 and GAD-7 scores were all positively and significantly correlated with mood stabilizers and stimulant prescriptions (Fig 4), Long COVID patients taking either class of medication had significantly higher GAD-7 scores compared to those not taking them (p = 0.031 and 0.035, respectively; Table 3), without significant differences in PCL-5 scores. These results persisted with the background of high variability of duration of prescription prior to study intake.

**Figure 4.**
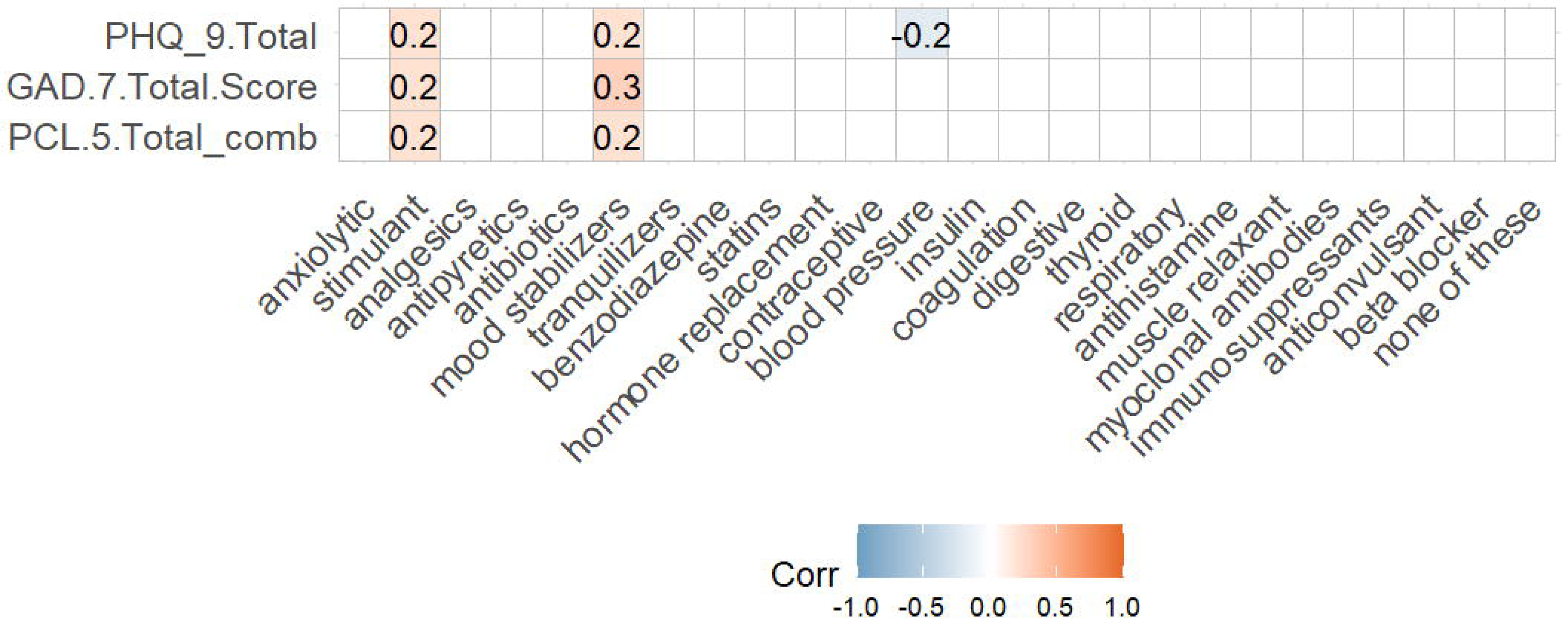
Correlation of depression (PHQ-9), anxiety (GAD-7), and stress (PCL-5) symptomatology with current medications. Spearman rank correlations were calculated for the intersection of {HQ-9, GAD-7, and PCL-5 total scores with classes of current medications endorsed by self-report. Correlations are given for all findings with *p* < 0.05.

**Table 3.** Correlation of anxiety, stress, and depression symptomatology with mood stabilizer and stimulant class medications.

| **Mood Stabilizer Class** |  |  |  |  |  |
| --- | --- | --- | --- | --- | --- |
| **Characteristic** | **Yes, N = 6<sup>1</sup>** | **No, N = 156<sup>1</sup>** | **Difference<sup>2</sup>** | **95% CI<sup>2,3</sup>** | **p-value<sup>2</sup>** |
| GAD-7 Score | 15.8 (6.3) | 8.2 (6.0) | 7.6 | 1.0, 14 | 0.031 |
| (Missing) | 0 | 19 |  |  |  |
| PCL-5 Score | 54 (23) | 30 (21) | 24 | -0.49, 48 | 0.053 |
| PHQ-9 Score | 18 (7) | 10 (7) | 7.9 | 0.89, 15 | 0.034 |
| (Missing) | 0 | 16 |  |  |  |
| **Stimulant Class** |  |  |  |  |  |
| **Characteristic** | **Yes, N = 8<sup>1</sup>** | **No, N = 154<sup>1</sup>** | **Difference<sup>2</sup>** | **95% CI<sup>2,3</sup>** | **p-value<sup>2</sup>** |
| GAD-7 Score | 13.1 (4.7) | 8.3 (6.1) | 4.9 | 0.46, 9.3 | 0.035 |
| (Missing) | 1 | 18 |  |  |  |
| PCL-5 Score | 48 (23) | 30 (21) | 18 | -1.7, 37 | 0.068 |
| PHQ-9 Score | 17 (5) | 10 (7) | 7.1 | 2.4, 12 | 0.009 |
| (Missing) | 1 | 15 |  |  |  |

<sup>1</sup> Mean (SD)
<sup>2</sup> Welch Two Sample t-test
<sup>3</sup> CI = Confidence Interval

## Discussion

Despite the best efforts of public health and medical systems, the COVID-19 pandemic remains a burden in much of the world, underscoring the need for further research into both the impact of acute infection and its associated infection-associated chronic condition, known as Long COVID (2–5).. SARS-CoV-2, the virus that causes COVID-19 and thus Long COVID, is neurotropic (24–27) and thus propagates throughout the central nervous system. The symptom clusters experienced by both acute COVID-19 and Long COVID patients imply significant distortions to normal functioning of central nervous system due to neurovirulent damage. In this study, we present key findings and acknowledge limitations, aiming to encourage others to build on this work to elucidate the underlying mechanisms of Long COVID and drive the development of effective therapies and interventions.

### Symptom Score is an effective clinical tool for Long COVID evaluation and longitudinal tracking

We characterized Long COVID symptoms in a cohort of patients who, due to the unique nature of our study site in South Texas, represented categories of patients thought to be a greatest risk(4,16,17,20) for severe COVID-19 infection and thus Long COVID (N = 162; 71% female, 53% Hispanic, median BMI = 30). Additionally, in contrast to previous studies (9,12,20), few patients with Long COVID symptoms in our cohort were ever hospitalized (N = 49) or required ventilation (N = 5). Thus, while our cohort uniquely highlights the relative share of Long COVID patients presenting for treatment in this region, we emphasize the disproportionate burden faced by Long COVID patients with similar demographic profiles across the United States.

We used the Symptom Score to correlate burden and complexity of multisystemic Long COVID symptomatology, which demonstrated neuropsychiatric symptom clusters that are highly consistent with larger and more detailed inventories of Long COVID symptoms from previous studies. Although relatively abbreviated, our findings align with attempts at deriving similar Symptom Scores in other large cohort studies (9,11,12) and reveal a high burden of fatigue, cognitive impairment, and anxiety among Long COVID patients, exacerbating the core Long COVID symptom profile. The significant impact on quality of life and the increased risk of worsening psychiatric disorders in Long COVID (28,29) underscore the need for greater attention to neuropsychiatric symptoms in the management of Long COVID. Thus, we advance the Long COVID Symptom Score as an effective inventory or core symptoms across several major systems (e.g., cognitive, respiratory, neuropsychiatric, pain, and constitutive/gastrointestinal symptoms) for use in clinical settings that benefit from rapid screening tools. We encourage clinicians and researchers to validate the Symptom Score in diverse populations. The Symptom Score may also assist with triaging patients for additional tests to detect neurovascular changes (30), neuroinflammation (31,32), and structural alterations (33–35) in patients with Long COVID.

### Psychiatric comorbidity is more predictive of Long COVID severity than patient factors

We hypothesized that chronic stress and mood disorders may significantly contribute to the complexity of Long COVID symptomatology, playing a key role in the abnormal neuropathophysiology associated with the condition. Due to the retrospective nature of this study, we are unable to assign causality to the pre-infection presence of neurocognitive, neurological, or neuropsychiatric symptoms to the Long COVID syndrome. Yet, our study is the first to administer a battery of validated scales to track with Long COVID symptom burden, resulting in strong, significant correlations across the board. Moreover, we assert that the neuropsychiatric profile of Long COVID patients is complex and merits refined clinical evaluation as was undertaken with this cohort.

We found that comorbid stress and mood disorder symptomatology as measured by the PHQ-9, PCL-5 and GAD-7 inventories was more predictive of Long COVID symptomatology than any patient factor considered in this study. It is admittedly difficult to attribute stress, anxiety, or satisfaction with life measures specifically to COVID-19 infection or chronic Long COVID symptomatology without knowledge of any index or inciting traumatic events. We anticipate this may be a difficult confounding factor to address, given the general stress and anxiety levels induced by the global COVID-19 pandemic at the time of study recruitment. Yet, the presence of complex neuropsychiatric symptomatology is undeniable, and the use of these instruments in Long COVID patient assessment is not a standard component of clinical assessment in this population. We also identified patients with premorbid anxiety disorders, who were not excluded from this study based on the original criteria, that may serve as a refined subgroup for future analyses on changes in anxiety levels attributable to neuropsychiatric burden in Long COVID.

Future studies should include validated screening tools to enable quantification and thus more objective assessment of neuropsychiatric burden in Long COVID patients. This may aid the tracking of symptom progression over time. One gap in this study was access to an inventory to characterize symptoms related to the patient-reported cognitive impairment reported by 73% of our cohort at time of assessment. A standardized and validated tool to assess specific features of cognitive impairments in this and other populations could point to underlying neurobiological pathways or circuitry that are particularly vulnerable to neurovirulence of COVID-19 infection or chronic damage related to Long COVID.

### Pharmacotherapy indexing remains a gap in establishing Long COVID causality

Not all patients with acute COVID-19 infection progress to develop Long COVID symptoms. While the cause of Long COVID syndrome following acute COVID-19 infection has yet to be established, there is some evidence that specific medication classes during acute phase confers elevated risk of Long COVID conversion (e.g., NSAIDs (36)). Nonetheless, we performed an exploratory analysis of patient-reported pharmacotherapy classes at study intake against scores on neuropsychiatric instruments since our results established a strong correlation to higher neuropsychiatric burden and Long COVID Symptom Score. Although we suspect our analysis ran against background variation in prescription initiation and duration prior to or concurrent with Long COVID onset, we found weak positive correlations with stimulant and mood stabilizing class prescriptions with higher neuropsychiatric battery scores. On the other hand, we found a weakly negative correlation with PHQ-9 scores and reported all-class blood pressure medications. We hope these results generate sufficient intrigue to merit study of pharmacotherapy as an index event for two motives: (1) to establish any additional risk or protective effects of pharmacotherapy classes for Long COVID symptom burden, or (2) to determine if management of neuropsychiatric comorbidities alleviates overall Long COVID burden.

### Some study interpretations limited due to study setting

The results of this study highlight considerations of stress and anxiety in Long COVID patients. Given the retrospective study criteria and nature of the data collection, we are unable to temporally resolve the onset of Long COVID symptomatology with any measure considered in this study, including neuropsychiatric symptom burden as measured with validated instruments and the impact of pharmacotherapy on the same scales. We can neither confirm nor assume that all patients would have started prescriptions before initial COVID-19 infection, nor at some point along the development of Long COVID symptomatology. The latter would particularly impact patients with the greatest duration of Long COVID symptoms, as it leaves the greatest chance and window for seeking symptomatic treatment with any of the medication classes analyzed. Importantly, we also cannot rule out the impact of greater socioeconomic stressors experienced by historically marginalized groups on Long COVID Symptom Score, nor any measure of stress and anxiety used here. Our results reflect patient status captured within a single visit to a Rehabilitation Medicine clinic and thus would likely benefit from long-term follow-up with increased sample size.

## Materials and Methods

This study was conducted under the approval of the Office of the Institutional Review Board at the University of Texas Health Science Center at San Antonio Long School of Medicine (protocol #20210194EX). Participants in this retrospective cohort study were men and women who were evaluated in a Rehabilitation Medicine outpatient clinic in South Texas from January 2020 to July 2021.

The study setting was an outpatient physiatry clinic in which each participant was evaluated, in a private room, by clinicians for Long COVID symptomatology and administered screening tools for stress, anxiety, and quality of life measures. The data was retrospectively collected directly from the medical chart in the University of Texas at San Antonio Health Science Center’s Electronic Medical Record (EPIC).

Responses were recorded directly into RedCap and then aggregated in a de-identified database for statistical analysis.

### Analytical Population

Eligible participants included patients over the age of 18 years old with a history of acute COVID-19 infection confirmed either by positive COVID-19 test (89%) or evaluation by a clinician for COVID-19 symptoms. Patients were seen at varying periods post-COVID-19 infection. Participants were ineligible if they were younger than 18 years old, did not have a history of acute COVID-19 infection, were evaluated for conditions apart from Long COVID, or were unable to read or speak in English to complete screening tools and clinical evaluation.

During the study period, 235 patients were clinically evaluated. In our analysis, we excluded patients who either did not complete the PTSD Checklist-Civilian Version 5 (PCL-5), Patient Health Questionnaire (PHQ-9), Generalized Anxiety Disorder Scale 7 (GAD-7), or Satisfaction with Life Scale (SWLS). Patients were not excluded from analysis if they were missing information on other patient factors or screening tools. We excluded these individuals from some sub-analyses (“Missing”), as noted throughout this report. The final analytical sample included 162 patients. These screening tools are outlined in detail below.

### Patient Factors

In addition to the above outlined measures, we also collected patient factors for use as covariates in this study (Table 1). Patient factors included biological sex, ethnicity/race, age, education level, body mass index (BMI), pre-existing psychiatric disorders, current medication list, type of health care institution, region, and employment status.

### Long COVID Symptom Score

We collected self-reported Long COVID symptomatology noted in the medical record from a single clinical evaluation of Long COVID symptoms (9,11) from all participants. Long COVID symptoms included: (1) positive COVID-19 test at the time of primary infection, (2) fatigue, (3) coughing or shortness of breath (SOB), (4) muscle or joint ache, (5) headache, (6) increased time spent in bed, (7) fever, (8) loss of smell, (9) loss of taste, (10) cognitive impairment, (11) insomnia or sleep disturbance, (12) weight loss, (13) diarrhea, (14) nausea or vomiting, (15) rash, (16) currently exercising. For our analysis, we then operationalized Long COVID symptoms and calculated a Symptom Score based on the total number of symptoms reported during clinical evaluation. We also evaluated factors thought to affect PASC symptom severity, such as the amount of elapsed time from Long COVID evaluation to initial acute COVID-19 infection, whether the patient was hospitalized, required ventilation or supplemental oxygen, or experienced cardiac damage because of primary infection.

### Stress and Posttraumatic Stress Disorder (PTSD) Assessment (PCL-5)

Posttraumatic stress disorder (PTSD) Check List for DSM-5 (PCL-5 (37,38)) was used to assess stress and trauma-related symptoms. The PCL-5 was adapted for the Diagnostic and Statistical Manual of Mental Disorders-Fifth Edition (DSM-5) and demonstrated strong reliability and validity (39). The PCL-5 is a 20-item inventory designed to gauge symptoms based on 4 major PTSD symptom clusters: Cluster B (The traumatic event is persistently experienced), Cluster C (Avoidance of trauma-related stimuli after trauma), Cluster D (Negative thoughts or feelings began or worsened after the trauma), Cluster E (Trama-related arousal and reactivity that began or worsened after the trauma. Participants rated the degree to which they experienced each symptom on a scale ranging from 1 (not at all) to 5 (extremely), with possible scores ranging from 17 to 85. Cut scores range from 30 to 60 depending on the variation and base rate of the disorder in the population and settings, and it is recommended to use higher cut-offs in populations with higher base rates, such as veterans, and lower cut scores with populations of lower base rates of PTSD (40). A total score of 31-33 suggests a diagnosis of PTSD and that the patient may benefit from PTSD treatment.

### Anxiety Assessment (GAD-7)

We used the Generalized Anxiety Disorder-7 (GAD-7 (41)) to evaluate patients for presence and severity of anxiety. The GAD-7 is a self-report seven item questionnaire assessing symptoms of generalized anxiety disorder that has been found to have validity as a measure of anxiety in the general population (42). In this study, the GAD-7 was altered to reflect symptomatology within the time frame of the past month to match the frame of reference for other screening tools used in this study. GAD-7 scores were used to assign patients into standard categories based on anxiety-related symptoms: 0-4 = minimal anxiety, 5-9 = mild anxiety, 10-14 = moderate anxiety, 15-21 = severe anxiety.

### Depression Assessment (PHQ-9)

We used the 9-item Patient Health Questionnaire (PHQ-9 (43)) to evaluate patients for presence and severity of depression. The PHQ-9 has a cut score of 6 that has been recommended for depression screening in primary care and a score of 10 or higher is used to detect symptoms of major depressive disorder (44).

### Satisfaction with Life Assessment

The Satisfaction with Life Scale (SWL (45)) is a five item self-report Likert scale (“strongly disagree” to “strongly agree”) to measure satisfaction with life as a proxy for subjective well-being. Past studies have established adequate reliability and predictive validity in a wide range of age groups (46). We analyzed patients who fell within categorical score ranges.

### Statistical Analysis

We used open-source statistical software (R version 4.3+, Vienna, Austria) to complete all statistical analyses in this study. Patient factors were tested for associations with Symptom Score by stratifying participants as above or below the median symptom score and using the Wilcoxon rank sum test (continuous variables) or Fisher’s exact test (categorical variables). Medication use for each category was used as a predictor within linear regression models for each outcome (PCL-5, GAD-7, PHQ-9, and Symptom Score). Spearman’s correlations were used to identify associations amongst these outcomes. All testing was two-sided (α = 0.05).

## Data Availability

All data produced in the present study are available upon reasonable request to the authors.

## Acknowledgements

This was a study completed by the interdisciplinary collaborations of the South Texas Psychiatry Practice-Based Research Network (PBRN). We would like to express our gratitude to the patients who entrusted our team with the data for this study. Our also extend our sincere gratitude to the many students who performed data collection and contributed to the development of poster presentations of this study (Anne Marie Wells, Richa Sinkre, Ashley Chakales, Sean Rumney, and Phillip Yang). Finally, we thank the South Texas Psychiatry PBRN staff, especially Kaitlyn Waxler, for their persistence and keeping the study organized.

## Funding

This study (JG, MG) was supported by the National Center for Advancing Translational Sciences, National Institutes of Health, through Grant UM1 TR004538. The training for AMW was supported by F30MH134482, T32R004545, STX-MSTP (T32GM113896 / T32GM145432), T32NS082145, R25NS089462. MVG was supported by a grant from the Agency for Healthcare Research and Quality (AHRQ). The content is solely the responsibility of the authors and does not necessarily represent the official views of the NIH.

## Compliance with Ethical Standards

The authors have no disclosures.

## Notes

### Competing Interest Statement

The authors have declared no competing interest.

### Author Declarations

This study was conducted under the approval of the Office of the Institutional Review Board at the University of Texas Health Science Center at San Antonio Long School of Medicine (protocol #20210194EX).

### Summary of Updates

Additional discussion of results following peer review.

