## Supplemental for "Assessing Neuropsychiatric Symptoms in Long COVID: A Retrospective Cohort Study from a South Texas Long COVID Clinic"

**Affiliations**:

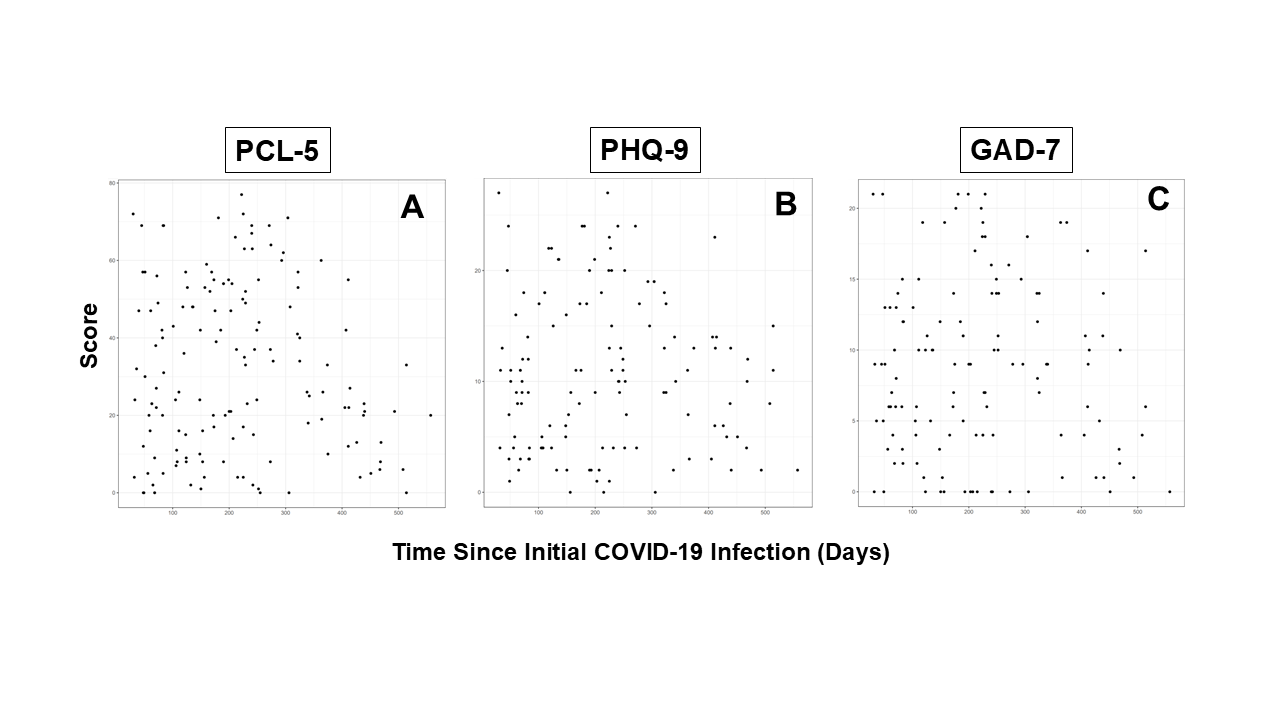

**Figure S1. Duration of COVID-19 symptoms is not correlated with patient scores on stress/PTSD, depression, or anxiety symptom screening tools.** Scatterplots of the correlation between time since initial COVID-19 infection (days) and: (A) stress/PTSD symptoms [PCL-5], (B) depression symptoms [PHQ-9], and (C) anxiety symptoms [GAD-7]. Screening tool scores (y-axis) are scaled relative to the score range of each inventory. There was no correlation time elapsed since initial infection any screening instrument for psychiatric symptoms used in this study.

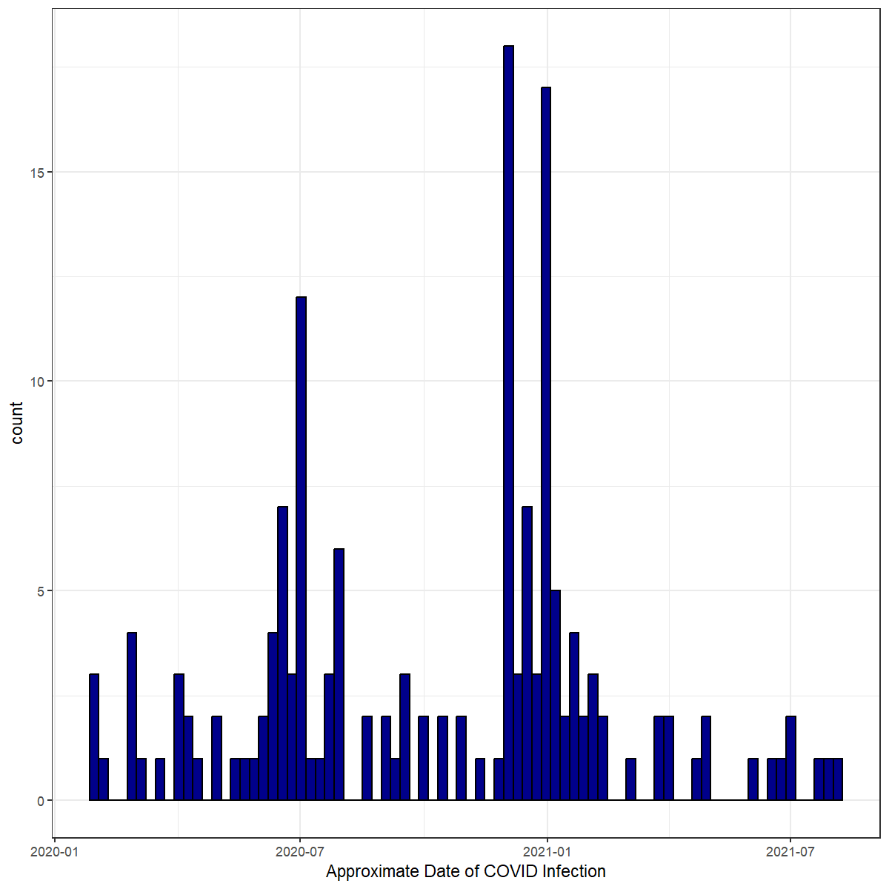

**Figure S2. Onset of initial COVID-19 infection in analytical sample aligns with major waves of pandemic.** We plotted the approximate date of initial COVID-19 infection in our patient sample and compared the distribution of peaks with data from local (Bexar County) and national (United States) peak rates of infection during the same time points.

**Table S1: Counts of current medications reported by patients at time of PASC evaluation.**

| **Characteristic** | **N = 162***^1^* |
| --- | --- |
| **respiratory** | 66 (41%) |
| **anxiolytic** | 55 (34%) |
| **analgesics** | 51 (31%) |
| **antihistamine** | 43 (27%) |
| **digestive** | 40 (25%) |
| **blood pressure** | 33 (20%) |
| **statins** | 25 (15%) |
| **none of these** | 19 (12%) |
| **thyroid** | 18 (11%) |
| **insulin** | 17 (10%) |
| **beta blocker** | 16 (9.9%) |
| **muscle relaxant** | 15 (9.3%) |
| **hormone replacement** | 14 (8.6%) |
| **anticonvulsant** | 11 (6.8%) |
| **coagulation** | 11 (6.8%) |
| **antipyretics** | 10 (6.2%) |
| **benzodiazepine** | 9 (5.6%) |
| **immunosuppressants** | 9 (5.6%) |
| **contraceptive** | 8 (4.9%) |
| **stimulant** | 8 (4.9%) |
| **antibiotics** | 7 (4.3%) |
| **mood stabilizers** | 6 (3.7%) |
| **tranquilizers** | 5 (3.1%) |
| **myoclonal antibodies** | 2 (1.2%) |
| **none of these** | 19 (12%) |
| *^1^* n (%) | |
